# Women’s and healthcare providers’ experiences of care during childbirth at public health facilities in a conflict-affected area of Nigeria

**DOI:** 10.1101/2025.04.09.25325543

**Authors:** Shatha Elnakib, Ayodeji Kazeem Olalekan, Charity Maina, Rejoice Helma Abimiku, Sussan Israel-Isah, Emilia Ngozi Iwu, Meighan Mary, George Odonye, Hannah Tappis

**Affiliations:** International Health Department, Johns Hopkins Bloomberg School of Public Health, Baltimore MD USA; Center for Humanitarian Health, Johns Hopkins University, Baltimore MD USA; Institute of Human Virology, Abuja, Nigeria; Rutgers University School of Nursing, Newark, NJ, USA; International Center for Maternal and Newborn Health, Johns Hopkins Bloomberg School of Public Health, Baltimore, MD, USA

**Keywords:** Nigeria, maternal health, quality of care, person-centered maternity care, conflict-affected settings

## Abstract

**Background:** Person-centered maternity care (PCMC) is an essential aspect of quality maternal healthcare. However, limited research has been conducted on PCMC in Northeast Nigeria, where the Boko Haram insurgency has impacted healthcare delivery. We sought to assess the status of PCMC as well as women and providers’ perceptions of safety and security in the most utilized public health facilities in Yobe State.

**Methods:** This cross-sectional study draws on data collected through 167 postpartum patient exit surveys and 32 surveys with maternity care providers in all public health facilities with an average of five or more births per day in Yobe State, Nigeria. Results were analyzed using descriptive statistics to describe the environment of care, and PCMC was calculated, using a validated 13-item scale. We calculated an overall PCMC score as well as scores for each of the three PCMC sub-domains. We additionally assessed bivariable associations between PCMC and several factors like facility type, women’s characteristics, and provider training in respectful care.

**Findings:** The overall PCMC score was not high (mean 27 of 39, 70% of the maximum score - standardizing scores to a 0-100). While respectful care and dignity (mean 9/12, 78%) and supportive care (mean 7/9, 78%) were rated relatively high, effective communication and autonomy scored notably lower (mean 10/18, 57%). Women reported challenges, including limited ability to ask questions (33%), lack of autonomy in choosing birthing positions (12%), and inadequate explanations of prescribed medications (12%). Women reporting household internal displacement, those with less education and those who delivered in higher level facilities tended to report worse PCMC scores. However, both providers and women generally reported feeling safe within healthcare facilities.

**Conclusion:** Poor communication between provider and patient and lack of support for women’s autonomy were important contributors to suboptimal PCMC in our study. Our findings highlight the vulnerability of internally displaced women, and those with less education to poor care and highlight key areas for targeted improvement and intervention.

## INTRODUCTION

While global maternal mortality has declined in the period 2000-2020, maternal mortality remains high in sub-Saharan African countries, with 70% of global maternal deaths occurring in the African region [1], [2]. This is in part due to poor quality of maternity care at health facilities [3]. A large body of evidence indicates that women experience high rates of disrespect and abuse, which has directed attention to the importance of person-centered maternity care (PCMC) in quality improvement efforts at facilities [4]. PCMC is defined as care that respects and responds to women’s expressed preferences, needs, and values, and ensures that women’s values guide all clinical decisions [5], [6]. This type of care elevates women’s autonomy and choices and reinforces their involvement in decision making around their care.

PCMC is important not only because it improves patient experience but also because it is associated with improved health outcomes [7]. Ensuring that care is respectful and responsive, can lead to improved satisfaction and engagement by focusing on individual patient needs, preferences, and values [8]. Allowing patients to take part in birth decision-making also fosters trust and effective communication between them and the health providers during childbirth, contributing to positive childbirth experiences [9].

Despite its importance in improving healthcare utilization and outcomes, PCMC remains poor in many settings [4]. Studies conducted in select urban and rural facilities in Kenya, Ghana, and India reveal significant gaps: in Kenya and India, just over half of interviewed women felt respected, while only 12% in Ghana reported the same [10]. Verbal abuse affected 11-18% of women, though physical abuse was rarer (<5%). Over 20% of women in Kenya and India were exposed during exams without coverage, and many, particularly in India, noted that providers never introduced themselves or used their names [10].

In Nigeria, the proportion of women who give birth in health facilities varies substantially, ranging from less than 10% to more than 80% of births depending on the state [11]. Over the last decade, national and state policymakers have documented commitments to improving quality of maternity care, including adoption of the Respectful Maternity Care Charter in 2013 and a national quality of care framework and strategy that places emphasis on person-centered care in 2018 [12]. However, progress in translating commitments into practice is slow, and studies from across the country have documented persistent dissatisfaction with the quality of maternity care [13] [14], [15]. Mistreatment of women during facility-based childbirth is widespread. A few Nigerian studies involving healthcare providers highlighted issues such as verbal and physical abuse, lack of visual privacy, denial of companionship, and restrictions on birth position [16], [17]. Factors hindering health workers’ ability to provide quality, PCMC include high workload, inadequate training, a focus on positive birth outcomes, and lack of incentives, while poor working conditions, such as inadequate infrastructure and stressful hospital protocols, serve as institutional drivers of disrespectful maternity care [18] [19].

To our knowledge, no studies have examined women’s experiences of PCMC in conflict-affected areas of Northeast Nigeria, despite conflict-related stressors that can exacerbate provider workloads, poor working conditions, and women’s vulnerability to ill-treatment. The objective of this study was to assess experiences of care during childbirth in the most utilized public health facilities in Yobe State in Northeast Nigeria. We additionally explored associated factors, investigating whether displacement status, parity, age, and level of facility impacted PCMC. To gain insight into the care environment, we also examined women’s and maternity care providers’ perceptions of the safety and security of public health facilities where births took place.

## METHODOLOGY

### Study design

The analysis presented in this paper is part of a larger cross-sectional assessment using multiple methods to assess quality of maternity care at high-caseload public health facilities in Yobe State in Northeastern Nigeria [20]. This analysis draws on data collected through exit surveys with postpartum women who had vaginal births at these facilities and through interviews with maternity care providers in the same sites. Data collection tools were adapted from the Demographic and Health Survey (DHS) Program Service Provision Assessment (2022 revision) tools [21] with additional content added to capture health worker experiences and considerations related to conflict-affected contexts (Supplementary Files 1 and 2).

### Study setting and participants

Yobe has an estimated population of approximately 3.4 million people [22]. The Boko Haram insurgency has led to the destruction of health facilities, displacement of healthcare workers, and disruption of maternal and newborn health services. Only about 32% of births in Yobe occur at health facilities, reflecting challenges in healthcare access exacerbated by the ongoing conflict [11].

All public health facilities in Yobe State averaging five or more births per day reported in the national health information system were included in the study. The threshold of at least five births a day was selected based on WHO Health Facility Assessment Guidelines for assessment of quality of care, and past experience conducting similar studies, as a pragmatic threshold to ensure that data collectors would be able to survey multiple women being discharged after childbirth on each day visiting a facility [23], [24]. In total, four out of 275 public facilities expected provide maternity services met this eligibility criteria. Three of these facilities were hospitals (two tertiary hospitals and one general hospital) with surgical capacity, and one was a primary health centre (PHC). All four facilities were located in urban areas: Damaturu – the state capital with an estimated population of 137,900, and Potiskum - one of the most populous and fastest-growing cities in Yobe State, with a population of 483,346 as of 2022 [25], [26]. Both Damaturu and Potiskum have faced significant security challenges, particularly during the peak of the Boko Haram insurgency that severely disrupted maternal and newborn health services. This included the destruction of healthcare facilities, displacement of staff, theft of medical equipment and increased risks of abduction of healthcare workers. However, efforts have been made to enhance security and return to normalcy.

Women and girls discharged with their newborns within 48 hours after a live birth at these facilities were invited to participate in structured postpartum exit surveys. Assuming a design effect of 1+ (*m* – 1)x*rho* using *m =* the average number of births per facility and *rho* = 0.1 correlation of quality-of-care outcomes within facility, we estimated that conducting structured postpartum exit surveys with 43 women per facility would allow us to assess a range of primary dichotomous outcomes. This sample size provides sufficient power to determine whether key international quality-of-care standards are met, with a margin of error of ±10 percentage points and a 95% confidence interval. As the PCMC scale incorporated in the DHS Service Provision Assessment has not been validated for cesarean birth, this analysis focuses on the subset of respondents discharged within 48 hours after a vaginal birth [21].

In addition, all maternity staff responsible for providing intrapartum care at these facilities were invited to participate in a structured survey to understand their knowledge, attitudes, practices, and constraints faced in service provision. Inclusion criteria for these interviews were broadened from skilled birth attendants on duty during the data collection period to maternity care providers to allow for inclusion of community health extension workers - trained healthcare providers in Nigeria who deliver primary healthcare services, and who are charged with attending births in the selected facilities [27] [28]. Note that in areas of Nigeria with healthcare worker shortages, it is common for community health extension workers based at primary health care centers to conduct deliveries even if they have not completed additional in-service training required to qualify as a skilled birth attendant [27].

### Data collection

Data collection was conducted as part of a larger assessment of quality of maternity care services at selected facilities conducted between February 2023 and April 2023. The assessment team comprised of nine data collectors with a backgrounds in the health profession (including both female nurse-midwives and male public health specialists), trained on the assessment tools, data collection and quality assurance procedures and research ethics. Data collectors visited facilities in teams of 4-5, one assigned to facilities in Damaturu and the other to facilities in Potiskum. Each team had at least one female to ensure participants had an option for speaking with a data collector of the same gender.

For postpartum exit interviews, maternity ward staff informed eligible patients about the study as they were preparing for discharge. Those willing to consider participating were then introduced to a study team member waiting outside of the postpartum ward/room and escorted to a private room for informed consent and interview. Maternity care providers were informed about the study at the beginning of the data collection period and invited to schedule an interview at the start or end of their shift to avoid interference with patient care duties.

Written informed consent was obtained prior to commencing data collection. Face-to-face structured surveys with individual patients and maternity providers were conducted in English or Hausa, depending on the respondent’s preference, using standardized, pretested questionnaires (Supplementary Files 1 and 2) configured on REDCap mobile data collection application on Android tablets [29]. The postpartum patient exit survey consisted of a structured questionnaire with close-ended questions designed to assess women’s experiences of maternity care. This included evaluating aspects such as respectful treatment, communication with providers, informed decision-making, and experience during labor and delivery. Additionally, a separate questionnaire was administered to maternity care providers, focusing on their knowledge, training, and experiences in delivering maternal health services. This provider-focused survey also comprised close-ended questions that explored training in person-centered care, familiarity with clinical guidelines, and challenges encountered in service provision. In both questionnaires, participants were asked to rate the perception of safety and security in facilities in relation to 5 statements. They were asked if they strongly agree, agree, disagree or strongly disagree to each statement (Box 1).

#### Box 1

Statements measuring perception of safety and security in the facility asked to postpartum women and health providers

|  |
| --- |
| I feel safe from crime and violence when I am at the facility. |
| I feel safe from crime and violence when traveling to and from work at the facility. |
| The area surrounding this health facility is marked by repeated violence. |
| The level of violence in this area has increased a lot in the last year. |
| The level of safety and security around this facility is similar to where I live. |

### Analysis

Stata version 18 statistical software was used to perform the analysis [30]

Person-centered maternity care was measured using the shortened PCMC scale, a summative score combining responses to 13 of the original 30-item PCMC scale [6]. The shortened scale has been found to have high content, construct, and criterion validity as well as high reliability [3]. The scale has three subdomains: dignity and respect (4 items), communication and autonomy (6 items), and supportive care (3 items). Each subdomain is measured through several items on a four-point response scale, that is 0-‘no, never’, 1-‘yes, a few times, 2-‘yes, most of the time’ and 3-‘yes, all the time’ for a total score ranging from a low of 0 to a high of 39. Scores were standardized on a scale 0-100, with lower scores indicating poorer PCMC.

#### Box 2

Items under the PCMC scale by subdomain

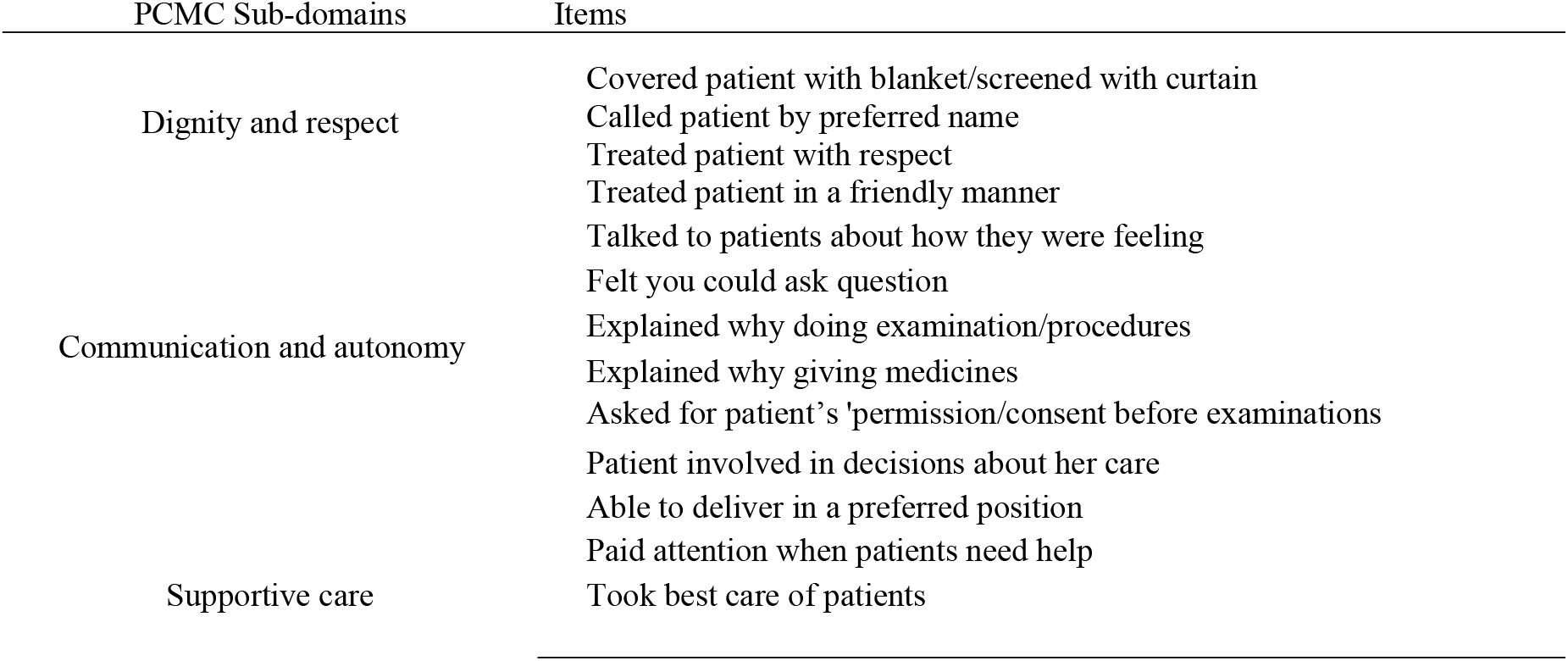

First, we assessed the normality of data under each of the three domains, using a histogram with normality curve, boxplot, and Shapiro wilk test of normality. Because the variables were not normally distributed, variables were standardized and an independent test was performed on the domains to examine whether there is a difference in women’s mean score of PCMC, dignity and respect, communication and autonomy, and supportive care across higher and lower levels of care. Higher levels of care denoted facilities that were secondary or tertiary care in nature. Lower levels of care denoted primary health facilities. We additionally investigated differences in PCMC across women’s age (dichotomized into those below age 25 years and women aged 25 years and older), education (none, primary, secondary/higher), parity (primipara those who had a birth (the day before), multiparous -women who have had 2 to 4 births before, grand multiparous -those who have had five births or more) and displacement status (whether respondent reported that she or her household experienced displacement vs not). We used t-tests and analysis of variance tests to determine if there are statistically significant differences between the PCMC means of the subgroups.

In addition to PCMC, we analyzed provider training in PCMC using descriptive statistics, specifically whether providers reported ever receiving any in-service training, training updates or refresher training on topics related to intrapartum (delivery) care and respectful maternity care in specific. We additionally analyzed variables that captured perceptions of safety and security of the care environment among women and health providers

We presented descriptive statistics of the five statements describing perceptions of safety and security in the sample and compared responses between women and health providers, and those in higher vs lower levels of care.

### Ethics approval

Ethical approval was obtained from the Johns Hopkins Bloomberg School of Public Health Institutional Review Board (IRB00021922) Yobe State Health Research Ethics Commission (MOH/GEN/747/VOL.I). All participants provided written informed consent.

## RESULTS

### Characteristics of women and maternity care providers

Table 1 describes participants’ background characteristics by facility type. Overall, 167 of the 186 (90%) postpartum women interviewed had vaginal births and are included in this analysis. The mean age of these women was 28 years with a standard deviation (SD) of 7 years. Approximately one-quarter (27%) had no education compared with 64% who had secondary or higher education. Thirteen percent of respondents were primiparous. Almost all participants (95%) planned to give birth at the facility where they were interviewed. One in four (24%) participants reported that they or members of their household experienced displacement from their homes for at least one month in the last year.

**Table 1:** Background characteristics of women by facility type.

| Variables | Total<br>n(%) | Primary<br>health<br>centre<br>n(%) | Tertiary/<br>secondary<br>hospital<br>n(%) | P-value |
| --- | --- | --- | --- | --- |
| Total | 167 (100.0) | 44 (100.0) | 123 (100.0) |  |
| **Women's age-group** |  |  |  | 0.35 |
| 15-19 years | 12 (7.2) | 4 (9.1) | 8 (6.5) |  |
| 20-24 years | 47 (28.1) | 12 (27.3) | 35 (28.5) |  |
| 25-29 years | 48 (28.7) | 8 (18.2) | 40 (32.5) |  |
| 30-34 years | 32 (19.2) | 11 (25.0) | 21 (17.1) |  |
| 35-39 years | 15 (9.0) | 3 (6.8) | 12 (9.8) |  |
| 40-44 years | 11 (6.6) | 5 (11.4) | 6 (4.9) |  |
| 45-49 years | 2 (1.2) | 1 (2.3) | 1 (0.8) |  |
| Mean (SD) | 27.7 (6.7) | 28.9 (7.9) | 27 (6.3) |  |
| Median (IQR) | 27 (10.0) | 28 (11.5) | 26 (10) |  |
| **Women's educational status** |  |  |  | 0.61 |
| No education | 43 (26.7) | 11 (26.2) | 32 (26.9) |  |
| Primary | 16 (9.9) | 3 (7.1) | 13 (10.9) |  |
| Secondary | 64 (39.7) | 20 (47.6) | 44 (37.0) |  |
| Higher | 38 (23.6) | 8 (19.0) | 30 (25.2) |  |
| **Parity** |  |  |  | 0.64 |
| 1 (Primipara) | 22 (13.2) | 4 (9.1) | 18 (14.6) |  |
| 2-4 births (Multipara) | 78 (46.7) | 21 (47.7) | 57 (46.3) |  |
| 5+ births (Grand multipara) | 67 (40.1) | 19 (43.2) | 48 (39.0) |  |
| Mean (SD) | 4.4 (2.9) | 4.9 (3.2) | 4 (2.7) |  |
| Median (IQR) | 4 (5) | 4 (5) | 3 (5) |  |
| **Marital status** |  |  |  |  |
| Yes, currently married | 167 (100.0) | 44 (100.0) | 123 (100.0) |  |
| **Planned to give birth at study site** |  |  |  | 0.22 |
| Yes | 159 (95.2) | 44 (100.0) | 115 (93.5) |  |
| No, sought care at facility after<br>having problem during home birth | 2 (1.2) | 0 (0.0) | 2 (1.6) |  |
| No, other reason | 6 (3.6) | 0 (0.0) | 6 (4.9) |  |
| **Participants and/or household**<br>**members displaced for 1 or more**<br>**months within past year** |  |  |  | <0.001 |
| Yes | 40 (23.9) | 0 (0.0) | 40 (35.8) |  |
| No | 127 (76.1) | 44 (100.0) | 83 (67.5) |  |

A total of 32 maternity care providers were interviewed across the four facilities, and most were females (88%), with 47% below 30 years of age. More than half of the providers were midwives or nurse midwives; only the PHC had community health extension workers involved in maternity care. Most providers had worked at no more than two different facilities before their current position. About 59% of healthcare providers have received in-service training on intrapartum or maternity care and of those, 68% had ever received in-service training on respectful maternity care (RMC, Table 2). Overall, providers at the PHC level were more likely to report receiving training (100%) compared to those working in secondary and tertiary hospitals (48%) (Table 2).

**Table 2:**
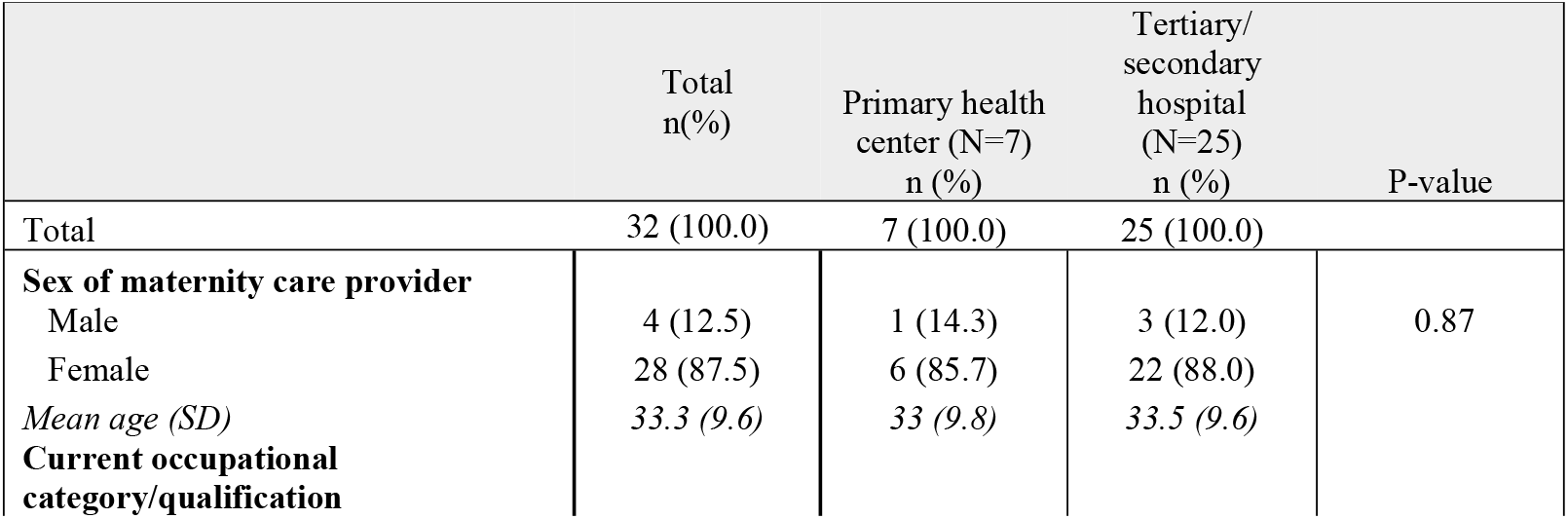
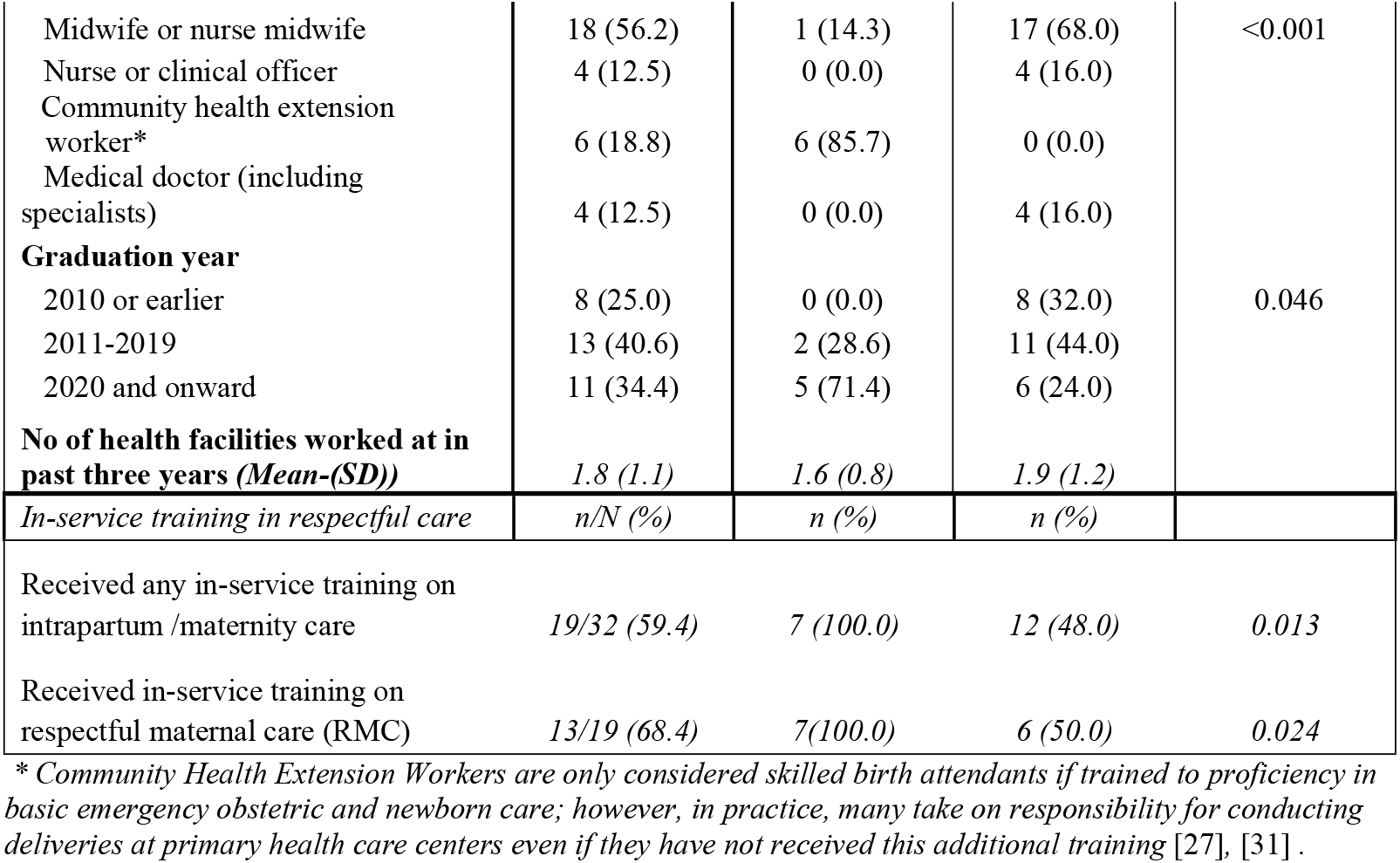
Health care provider characteristics and in-service training received by facility type.

***Figure 1: Responses to person-centered maternity care questions among women who delivered at a health facility***

***\*Numbers do not add to 100 because we are not depicting the percentage of participants who answered “I don’t know”***

### Person-centered maternity care scores among women

Table 3 presents the summary scores for the PCMC, with a maximum possible score of 39 based on responses to 13 items. The findings are organized by domain, each with its own maximum score and corresponding set of items. The mean PCMC score was 27 – equivalent to an average of 70% of the maximum possible score – and standard deviation of 6.0 reflecting high heterogeneity in responses.

**Table 3:** PCMC Scores among women and training in PCMC among maternity care providers.

| Quality of Care Variables | Total (mean score, SD) | Facility Type |  |
| --- | --- | --- | --- |
|  |  | Primary health centre (Lower) (mean score, SD) | Tertiary/ secondary hospital (higher) (mean score, SD) |
| *Women’s perception of quality of care* |  |  |  |
| Effective communication and autonomy (max score=18) | *10.3, 3.8* | *10.4, 5.2* | *10.3, 3.2* |
| Respect and dignity (max score=12) | *9.4, 2.0* | *10.9, 2.0* | *8.9, 1.7* |
| Supportive care (max score=9) | *7, 1.7* | *8.5, 1.3* | *6.5, 1.6* |
| PCMC (max score=39) | *26.8, 5.6* | *29.7, 7.2* | *25.7, 5.1* |

\*SD=Standard Deviation
*Effective communication and autonomy*

Six items reflected this domain with a maximum score of 18 points. The mean score for this domain was 10.3, equivalent to 57%, and the SD was 3.8. In the assessment of the communication domain, respondents indicated varying experiences, with some replying “most of the time” to questions related to effective communication practices. However, a third (33%) responded that they could never ask questions, 12% responded that they were not able to be in the position of their choice, and 12% said that doctors did not explain to them why they were giving them medicines.

#### Respectful care and dignity

Four items reflected this domain with a maximum score of 12 points. The mean score for this domain was 9.4, with SD of 2.0, equivalent to 78%. Most participants consistently responded, “all the time” and “most of the time” to items related to respect, including whether providers called them by name, treated them with respect, interacted with them in a friendly manner, and covered them with cloth or screened them with a curtain to prevent exposure.

#### Supportive care

Three items reflected this domain with a maximum score of 9. The average score in this domain was 7, equivalent to 78%. For this domain also, most of the participants consistently responded that they received supportive care “all the time” or “most of the time,” operationalized as i.) a provider paid attention when they needed help, ii.) a provider talked to them about how they were feeling, and iii.) a provider took the best care of them.

The findings from each of these domains indicate that while participants generally rated Respectful Care and Dignity and Supportive Care domains relatively high, scores on communication and autonomy were substantially lower.

### Variation in PCMC scores by individual characteristics and facility type

We found a statistically significant mean difference in PCMC between women who gave birth in the PHC vs those who gave birth in the three hospitals, with a mean difference of 4 points on the 39-point scale or 10% (p < 0.001). Women who delivered at the PHC experienced higher PCMC scores across domains compared to those who delivered at higher-level facilities. There was a statistically significant difference in the mean scores for respect and dignity, as well as supportive care, with women who received care at the PHC faring better compared to those who gave birth at one of the hospitals. However, no significant mean difference was observed in effective communication based on the level of facility where women gave birth.

The relationship between women’s experience of displacement (for themselves or a member of their household) and PCMC was also assessed. The findings revealed a significant mean difference in PCMC scores between women who reported displacement and those who did not (p<0.001). Additionally, women who received primary or secondary and higher education were more likely to report high PCMC compared to women with no education (p=0.026 and 0.002 respectively). However, PCMC scores did not differ significantly between younger and older women or between primipara and multiparous women. Detailed results are presented in Table 4.

**Table 4:** Mean differences in person-centered maternity care scores by facility and individual characteristics.

| Characteristic | Mean difference (95% CI) | P-value |
| --- | --- | --- |
| **Facility level (reference, PHC)** | R |  |
| Tertiary/secondary hospital | - 4.01(2.02, 6.0) | 0.0001 |
| **Displacement (reference, displacement)** | R |  |
| Did not report displacement for herself or member of the household | 4.70 (2.67,6.73) | <0.0001 |
| **Parity (reference, primiparous)** | R |  |
| Multiparous | -1.71 (-4.57, 1.16) | 0.241 |
| Grand Multiparous | -1.30 (-4.21, 1.61) | 0.380 |
| **Women's age group (reference, younger<25)** | R |  |
| Older (25+) | 0.98 (-2.90-0.93) | 0.312 |
| **Women's education status (reference, no education)** | R |  |
| Primary | 3.84 (0.46, 7.21) | 0.026 |
| Secondary/Higher | 3.28 (-4.63, 5.35) | 0.002 |

### Perception of women and maternity care providers on the safety and security of delivery facility

Approximately 99% of women strongly agreed or agreed that they felt secure from crime and violence within the health facility, compared to 88% of maternity care providers. Similarly, regarding perceptions of safety and security while traveling to and from the facility, 98% of women felt secure, in contrast to only 84% of providers (Table 5).

**Table 5:** Perceived safety and security level of the facility from postpartum women’s and maternity care provider’s perspectives.

| Indicators | Postpartum women |  |  |  | Maternity care provider |  |  |  |
| --- | --- | --- | --- | --- | --- | --- | --- | --- |
|  | Total | Strongly agree/ Agree | Strongly disagree/ Disagree | Don't know/Refused to answer | Total | Strongly agree/ Agree | Strongly disagree/Disagree | Don't know/ Refused to answer |
| Feel safe from crime and violence when inside this HF | 167 | 165 (98.8%) | 1 (0.6%) | 1 (0.6%) | 32 | 28 (87.5%) | 3 (9.4%) | 1 (3.1%) |
| Feel safe from crime and violence when traveling to/from this HF | 167 | 164 (98.2%) | 1 (0.6%) | 1 (0.2%) | 32 | 27 (84.4%) | 4 (12.5%) | 1 (3.1%) |
| The area surrounding this health facility is safe and peaceful overall | 167 | 163 (97.8%) | 0 (0.0%) | 4 (2.4%) | N/A | N/A | N/A | N/A |
| The area surrounding this HF is marked by repeated violence | 167 | 1 (0.6%) | 160 (95.8%) | 6 (3.6%) | 32 | 3 (9.4%) | 27 (84.4%) | 2 (6.2%) |
| The level of violence in this area has increased a lot in the last year | 167 | 0 (0.0%) | 163 (97.6%) | 4 (2.4%) | 32 | 1 (3.1%) | 30 (93.8%) | 1 (3.1%) |
| Level of safety & security around HF is similar to where women live | 167 | 152 (91.0%) | 11 (6.6%) | 4 (2.4%) | 32 | 30 (93.7%) | 2 (6.3%) | 0 (0.0%) |

The study further revealed that most respondents, both women and providers (98% vs. 94%, respectively), strongly disagreed or disagreed that the level of violence in the area where the facility was located had increased significantly over the past year. Similarly, a comparable proportion of women and providers (91% vs. 94%, respectively) believed that the level of safety and security around the facility was consistent with the level of safety in their residential areas.

When comparing perceptions of safety and security between women who gave birth at the PHC and the hospitals included in this study, a slight difference was observed (Table 5). Women reported higher perceived safety and security at PHC compared to hospitals; a similar pattern was found among maternity care providers.

Nonetheless, feelings of safety and security were not universal. Some maternity care providers at hospitals strongly disagreed with the statement that they feel safe from crime and violence within the facility (12%) and during travel to and from the facility (16%) and some reported that the area surrounding the facility was marked by repeated violence (12%) (Table 5). However, no statistically significant differences were identified for these safety and security indicators by facility level.

## DISCUSSION

Our study sought to assess women’s perceptions of PCMC at high-caseload public health facilities in Northeast Nigeria, a region where conflict and violent insecurity have severely impacted healthcare delivery. Overall, participants did not consistently receive high PCMC during childbirth, as reflected in an average PCMC score of 70 out of 100, indicating gaps in the provision of respectful, individualized, and responsive care. The subdomains of communication and autonomy received notably low scores, signaling important shortcomings with ensuring the meaningful engagement and involvement of women in care decisions. In contrast, the respect and dignity and supportive care subdomains scored higher, though many areas require attention and improvement. PCMC scores were significantly higher among women who delivered in the only PHC in Yobe State with an average of at least five births per day compared to those who delivered in hospitals with similar delivery caseloads. We identified significant disparities in PCMC based on education and displacement status, highlighting the heightened vulnerability of displaced and lower income individuals to suboptimal care. Further, our findings suggest that both providers and women generally feel safe within healthcare facilities, with women reporting slightly higher levels of safety than providers.

Our findings on the status of PCMC align with other studies, which highlight gaps in the quality of care [32], [33] [8], [34], [35]. A review examining the extent of PCMC during childbirth in low- and middle-income countries (LMICs) found that mean PCMC scores varied by region, with generally lower scores observed in Africa (below 75), moderate scores in Asia (ranging from 60 to over 90), and comparatively higher scores in the United States (above 80) [36].

There is an urgent need to ensure that women receive care that is responsive to their needs, as negative experiences with maternal care can deter women from seeking care altogether. Indeed, a study in Nigeria identified the negative attitudes of healthcare providers as a major barrier to seeking care among women [37] [36]. As in our study, a study conducted in Enugu State, in the southeast of Nigeria, found that the communication and autonomy domain tended to receive the lowest scores [8]. This was attributed to limited consented care and, similar to our study, inadequate explanations of medicines and low involvement of women in decisions about their care. The finding that 33% of women in our study reported feeling like they could not ask questions of their providers highlights the importance of fostering an environment where women feel empowered to actively participate in their care. Additionally, the finding that more than one-tenth of women could not request to be in the position of their choice during childbirth or did not understand the medicines being administered to them underscores the critical need to enhance communication strategies and ensure that care practices prioritize women’s autonomy and informed decision-making in these facilities. Several interventions evaluated in other settings have shown promise in enhancing communication between patients and providers. For example, communication skills training for midwives and doctors has been found to improve the quality of the information they give to women. Other interventions include offering women individualized information and giving women their own maternity records (referred to as women-held maternity records), with women who held their own records showing improvement in several aspects of communication [38].

Our scores for the respect and dignity and supportive care sub-domains were higher compared to other studies conducted in Nigeria [39], [40], [41], [42]. One study in Kano, in Northwest Nigeria found prevalence of obstetric violence to be high at 32% with almost 40% of women reporting neglect and abandonment and 23% reporting physical abuse [43]. In another review of studies examining obstetric violence in Nigeria, non-dignified care in the form of negative, poor and unfriendly provider attitude was frequently reported [42]. In contrast, in our sample most women felt like they were treated with respect and in a friendly manner all or most of the time.

Notably, women who gave birth in the urban, high-caseload PHC tended to have more positive experiences compared to their counterparts who gave birth in nearby public hospitals with similar or greater caseloads. This aligns with studies showing that maternity care quality often varies by facility level, with lower-level facilities often providing better person-centered treatment and more respectful care [44], [45]. This may be explained by the fact that lower-level care tends to be more individualized and person-focused, due to a confluence of factors that include lower patient loads, closer community ties, and greater provider-patient interaction. Our findings that providers in the PHC tended to report more frequent training in respectful maternity care and counselling and bereavement support could additionally explain the disparity in patient experience. In Nigeria, the National Primary Health Care Development Agency (NPHCDA) is the parastatal body responsible for supporting the planning, management and implementation of primary healthcare while higher-level facilities are under the authority of the state or Federal Ministry of Health [46], [47]. NPHCDA’s investments in training providers is an important step in improving the responsiveness of services to women’s needs and preferences, and evidence suggests that when done well, trainings and educational workshops can improve empathy and communication skills among healthcare professionals [48], [49], [50], [51]. Still, while training is one potential determinant of quality care, poor communication and limited autonomy during childbirth have been shown to stem from other factors such as high workload, inadequate staffing, and hierarchical cultures that prioritize medical decisions over patient preferences [52], [53], [54].

The finding that women who reported displacement in their household tended to have worse PCMC scores than their non-displaced counterparts warrant attention. This finding is consistent with other studies that find displacement status to be associated with disrespect and mistreatment during childbirth [55]. While studies exploring the experiences of Nigerian displaced persons are scant, studies in other LMICs [56], [57] highlight the differential treatment of women during pregnancy, labor, childbirth and the postpartum period based on social categories of vulnerability. Women who are less educated or who are displaced are often deprived of social support networks, experience increased poverty, and are consequently at a heightened risk of encountering discriminatory and stigmatizing treatment from healthcare providers [58] [59], [60]. Our findings underscore the importance of recognizing and addressing the specific challenges faced by internally displaced and lower income women.

Finally, our findings that both maternity care providers and postpartum women tended to report low levels of perceived insecurity in facilities and on the way to facilities is a positive one and underscores the importance of continuing to protect health facilities and providers. The Yobe State government has taken significant steps to enhance healthcare security and delivery. At the time data collection for our study began, government efforts were underway to rehabilitate and equip damaged health facilities through international partnerships, training and deploying healthcare workers, and constructing or upgrading 138 PHCs to improve access and safety [61], [62] [63], [64].

Our study holds significant implications for policy and programming, particularly in improving the quality of maternity care in Yobe State. Our findings underscore the pressing need to enhance PCMC in healthcare facilities, with a particular focus on ensuring that the perspectives, needs, and preferences of women are fully integrated into the design and delivery of care. This requires the development of mechanisms that systematically gather and incorporate women’s feedback, as well as the establishment of robust accountability frameworks that hold healthcare providers accountable. In Yobe State, a Quality Improvement Committee, which is a senior management body, oversees quality initiatives and quality of care. Opportunities to leverage the committee to advance person-centered care include integrating PCMC metrics into routine service quality assessments, ensuring that women’s voices are represented, and strengthening provider training on respectful and dignified maternity care.

Additionally, our study highlights the importance of designing and implementing interventions that are specifically tailored to meet the unique needs of vulnerable women, such as those of low socioeconomic status and internally displaced women. These targeted interventions should address the specific challenges faced by these women, including the erosion of social support networks, heightened security risks, and increased exposure to discriminatory attitudes from healthcare providers [65]. By addressing these gaps, policies and programs can significantly improve maternal health outcomes and foster a more inclusive, responsive healthcare system for all women, particularly those in vulnerable and marginalized positions.

Although our study focused on only four of the 275 public health facilities providing maternity services in Yobe, these four facilities accounted for nearly 20% of the facility births (12,771 of 66,484 births) reported in the state’s health information system in the year preceding the assessment and thus provides valuable insights into the quality of care available to women with facility births. Nevertheless, because data collection was limited to these four sites, all of which are located in urban centers, caution should be exercised in extrapolating findings to other lower-caseload PHCs and hospitals across Yobe State. Our study findings should also be interpreted in light of several additional limitations. First, our assessment of PCMC was based on the subjective perceptions of women, who may have low expectations of the health system and may normalize disrespect, abuse, and insecurity, particularly those who had healthy newborns and did not experience pregnancy or birth complications. Second, our inclusion criteria of facilities with an average of five or more births per day may have unintentionally limited our sample to facilities in safer locations, and as such, perceptions of care could differ in other, less secure facilities. Finally, we cannot rule out the possibility of social desirability bias and other biases that may have influenced women’s responses to our questions.

## CONCLUSION

Our study contributes to the scarce body of evidence documenting women’s experiences with healthcare in conflict-affected settings, offering valuable insights into the experiences of women with childbirth. Specifically, our findings emphasize the critical need to strengthen communication practices within healthcare settings, particularly with respect to ensuring that women are actively involved in decisions about their care. Furthermore, our study sheds light on the significant disparities in the quality of care between women receiving services at lower-level facilities and those attending higher-level hospitals, as well as the differences in care experienced by displaced versus non-displaced women. Addressing these issues is crucial to promoting positive childbirth experiences, reducing maternal and neonatal morbidity, and ultimately improving the overall healthcare outcomes for women in conflict settings.

## Data Availability

The data will be posted on Figshare

NA

## List of abbreviations

(PCMC): Person-centered maternity care
(DHS): Demographic and Health Survey
(RMC): Respectful maternity care
(PHC): Primary Healthcare
(NPHCDA): National Primary Health Care Development Agency

## Declarations

### Ethics approval and consent to participate

Ethical approval was obtained from the Johns Hopkins Bloomberg School of Public Health Institutional Review Board (IRB00021922) Yobe State Health Research Ethics Commission (MOH/GEN/747/VOL.I).

### Consent for publication

Not applicable

### Availability of data and materials

The de-identified dataset will be posted on Figshare

### Authors’ contributions

EI, CM and HT supervised the study. KA and SE conducted data analysis and led manuscript writing. CM, KA, GO, RA and SI contributed to data analysis and write-up of study findings. AK and GO supported data management and quality assurance. HT, EI, CM, and MM conceptualized the study. All authors have contributed to interpretation of the data, manuscript writing, and have read and approved the final version of the manuscript.

## Acknowledgements

We would like to acknowledge the data collection team and the women and providers who participated in the study.

